# 3D Printed N95 Equivalent for PPE Shortages: The Kansas City Mask

**DOI:** 10.1101/2020.08.23.20180513

**Authors:** Shiv Dalla, Brandon Bacon, Jack Ayres, Stephen Holmstead, Alan J. Ahlberg Elliot

**Affiliations:** University of Kansas Medical Center; University of Missouri—Kansas City, Truman Medical Center; Unaffiliated

## Abstract

**Introduction:** During the COVID-19 pandemic, the shortage of personal protective equipment (PPE) was well-reported and discussed, not only in the healthcare sector but across all of society as the demands for PPE skyrocketed. As hospitalizations for COVID-19-related illness continue to increase, many recent reports indicate the supply of PPE is persistently and significantly less than the demand. These PPE shortages encouraged communities of 3D printing experts and hobbyists to design and distribute homemade, 3D-printed PPE, including N95 mask substitutes. The mask presented, the Kansas City Mask (KC Mask), is one such product which was created from the maker community in partnership with local physicians and hospitals. This report discusses the design, manufacturing, and validation of the KC Mask design and its usage in the COVID-19 pandemic as well as future use as stopgap PPE.

**Methods:** The KC Mask was adapted from a similar design called the Montana Mask. Mask components were 3D printed and assembled then fit tested by qualitative fit testing (QLFT) at Truman Medical Center in Kansas City, MO as a proof of concept.

**Results:** The QLFT was successful and the KC Mask was approved for use by pandemic response administration staff at the hospital. Fortunately, the KC Mask has not required wide utilization, however, because supply chains for Kansas City area hospitals have, at the time of this publication, not yet been exhausted by the pandemic.

**Conclusion:** The results of Truman Medical Center’s approval of the KC Mask are promising for this N95 stop-gap substitute. Although further analysis and study is needed for this design, persistently increasing caseloads and PPE shortages necessitate an urgent dissemination of these preliminary results. The authors do not advocate for the KC Mask as a replacement of traditional N95 masks or other PPE but do endorse the KC Mask as a stopgap measure, proven to be effective in situations of dire PPE shortage based on CDC guidelines.

## INTRODUCTION

### Background

As the first eight months of 2020 have shown, the novel coronavirus, named “SARS-CoV-2,” represents a critical and serious threat to public health.^1^ Highly contagious and virulent, the virus has been reported to cause severe respiratory problems in infected patients and was declared a pandemic by the World Health Organization (WHO) in March.^1^ As the potential global economic impact of the pandemic came to light,^2^ the healthcare industry faced an overwhelming challenge: the adequate supply of appropriate personal protective equipment (PPE).^3^

### PPE Shortages

The shortage of PPE is well-reported and discussed, not only in the healthcare sector, but across all of society as the demands for PPE skyrocketed.^3-5^ Unfortunately, as hospitalizations for COVID-19-related illness continue to increase, recent reports indicate the supply of PPE is persistently and significantly less than demand.^6^ Even though the supply has risen since the beginning of the pandemic, experts predict that the supply is insufficient for predicted caseloads and it may take years in order for stockpiles to be fully replenished.^7^ PPE shortages have been shown to contribute to viral spread within healthcare environments and in the general community. This, of course, leads to increased societal harm and ultimately, preventable death. Of concern, these rates are shown to be significantly greater in communities of color and low socio-economic status.^8^3D printing experts and hobbyists, known as “makers” identified the challenge posed by PPE shortages and sought to create homemade masks, face shields and even gowns to mitigate the issue. Makers with access to 3D printers were encouraged to produce stop-gap PPE that could be used in clinical environments. The widespread availability and cost-effective nature of 3D printing lead to an incredible phenomenon of collaboration among makers to develop various PPE products. The flexibility and robust applications of 3D printers was leveraged into a uniquely useful tool to this end. Face shields, face masks, and strap adjustment devices became particularly popular products.

The mask presented, the Kansas City Mask (KC Mask), is one such product which was created from the maker community in partnership with local physicians and hospitals. This report discusses the design, manufacturing, and validation of the Kansas City mask design and its usage in the COVID-19 pandemic as well as future usage as stopgap PPE.

## METHODS

### Mask Design

The Kansas City Mask is a stop-gap, reusable, face mask substitute for level 1, level 3, or N95 face masks consisting of two 3D printed parts.

As shown in Figure 1, the main component of the mask is the contoured surface that makes contact with the user’s face. On the external surface of the main component there is a 60mm x 60mm square hole where the second component, the filter holder, a 60mm x 60mm grid, will snap into place, holding in the space filter material. The mask accepts most fabrics as filter material. In this study, Halyard H600 sterilization wrap, which is commonly used in hospitals for packaging of sterilized surgical instruments and N99 rated, was used.

**Figure 1:**
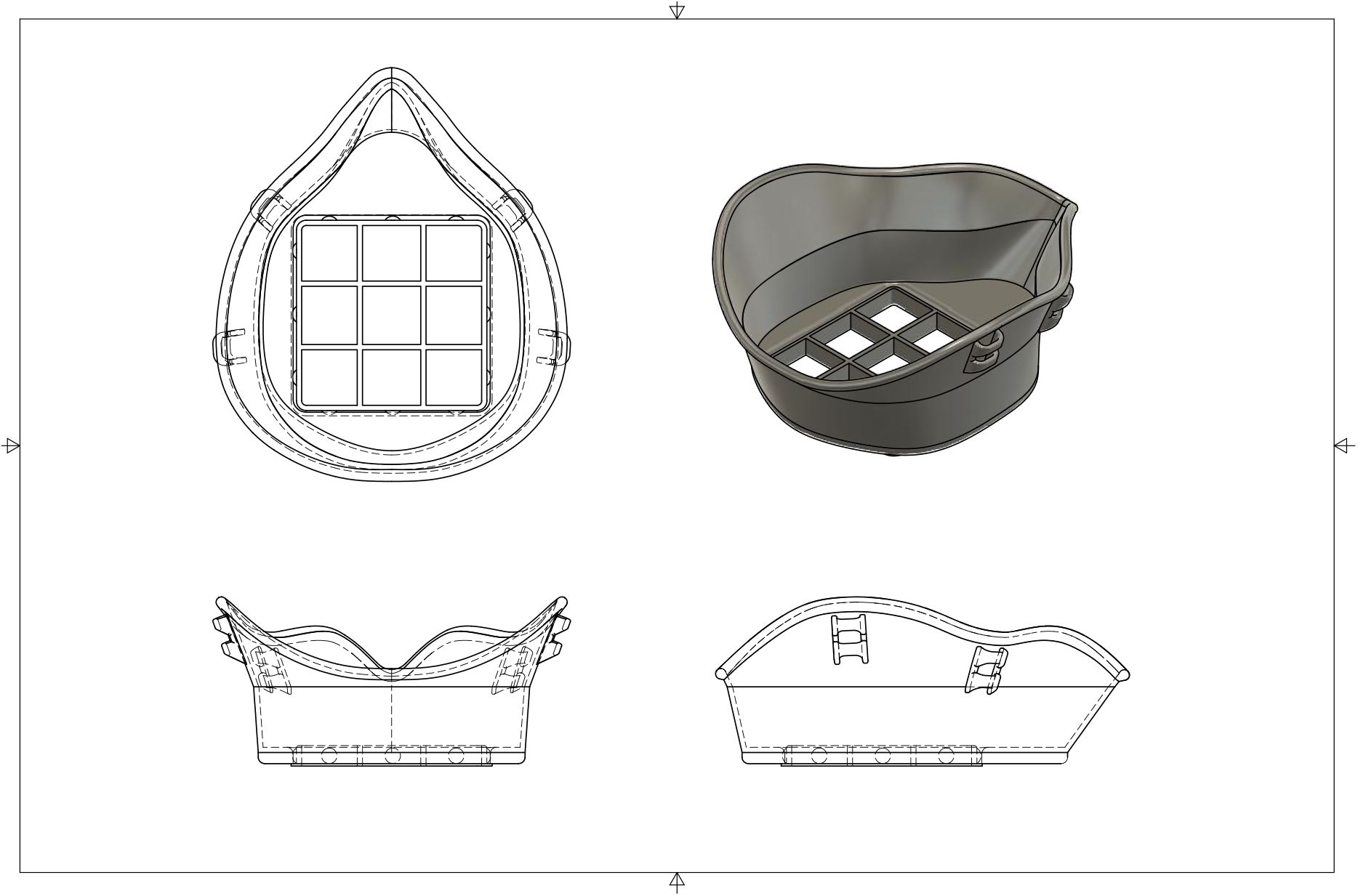
Multi view engineering drawing of KC Mask with 95 filter holder in place.

The Kansas City Mask was adapted from a similar design called the Montana Mask.^9^ The goal of this redesign was to address some shortcomings of the Montana Mask,^9^ namely ease of breathing and fit. The Montana Mask has a smaller filter cross sectional area, making it more difficult to breath. The design is additionally enhanced by the dipping process explained in its respective section.

### Design and 3D Printing

The design for the KC Mask was generated using commercial CAD software and exported to an STL file for compatibility with hobbyist 3D printer workflows. These workflows use a “slicer” software to convert the universal STL file into GCODE that is specific to an individual 3D printer. Two combinations of slicer and printer were tested. The Cura slicer was used to prepare GCODE for a Lulzbot Taz5, and PrusaSlicer was used to prepare GCODE for a Creality CR-10S. Masks were printed in a variety of filaments including Polylactic acid (PLA), Polyethylene terephthalate glycol (PET-G), and Thermoplastic polyurethane (TPU). PLA was selected for further fit testing for its ease of use, cost effectiveness, and ability to be safely thermoformed to a user’s face in hot water. For masks submitted to QLFT, the print settings shown in Figure 2 were used.

**Figure 2:**
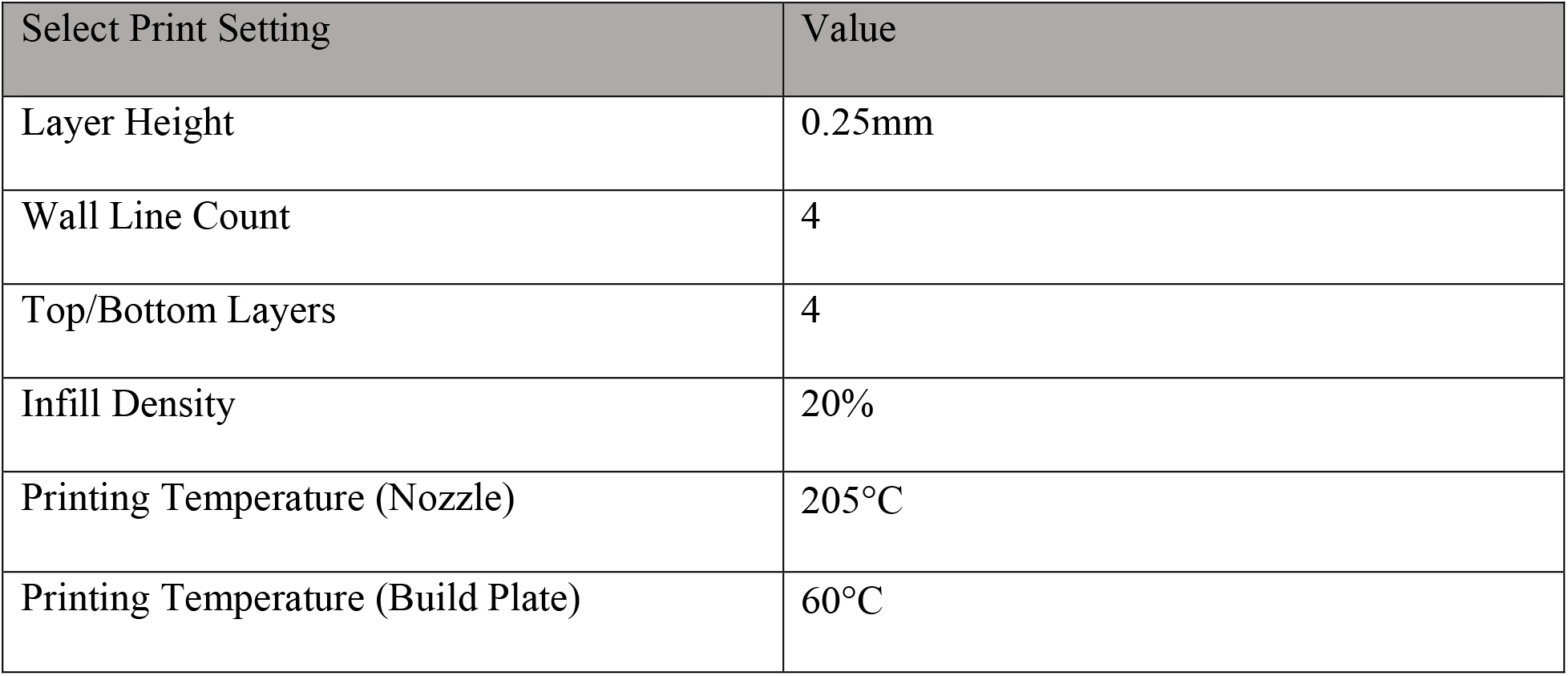
Print Settings.

### Dipping and Fitting

Dipping is an optional process in which liquid rubber is added to the rim of the mask which makes contact with the wearer’s face. This allows for a better seal as well as increased comfort. FlexSeal rubberized sealant was used in testing due to its lack of respiratory hazards. An optional dipping tray model is provided to help with this process. The goal is to coat the rim of the mask that will be touching the users face. Alternatively, weather stripping can be utilized for the same purpose, though this reduces ease of sterilization once applied.

After the mask is dipped, it can be molded to the wearer’s face by increasing the temperature of the mask above the glass transition point of the material. For PLA, the easiest way to do this is to submerge the mask (including the filter holder, but without any filter material) in hot water (~60°C). This allows for the PLA to become soft enough to mold, but the structure is maintained. The wearer should submerge the PLA for ~10 seconds and mold it to fit their face. This can be repeated until a good and comfortable fit is achieved.

### Fit Testing

The KC Mask was fit tested at Truman Medical Center in Kansas City, MO with the help of resident physician Dr. Brandon Bacon and the facilities staff. A standard qualitative fit test (QLFT) was performed using standard saccharin solution aerosol protocol.

### Usage

Elastic straps are used to secure the mask to the wearer’s face. In Figure 3, instructions for assembly and use of the mask are discussed. This image is distributed alongside the mask as instructions for the user. These instructions include how to mold the mask, place the straps, and insert the filter. Sterilization of the mask is done using a Sani wipe or dilute bleach solution. Filters are meant to be used only once, or can be re-sterilized per institutional protocol.

**Figure 3:**
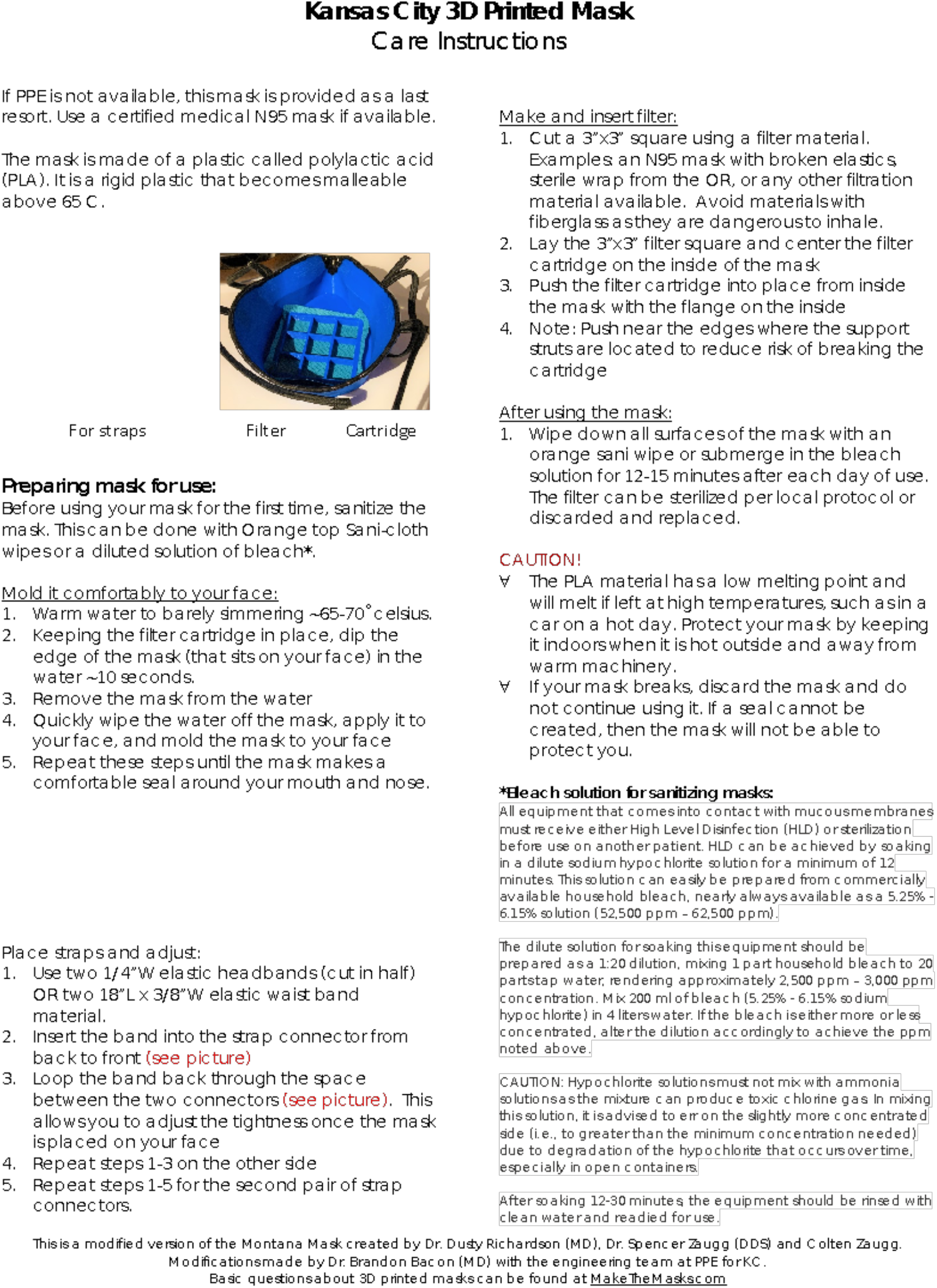
Assembly and Usage Instructions.

### Study Design

IRB approval was not requested nor required for this study because this was a proof of concept and quality improvement initiative. Although further analysis and study to prove efficacy is required, the purpose of this study and report is to discuss the design, manufacturing, and validation of the KC Mask concept.

## RESULTS

Several dozen masks were distributed to Truman Medical Center. As a proof of concept, Dr. Brandon Bacon donned the mask and performed a standard qualitative fit test (QLFT). The QLFT was successful and the KC Mask was approved for usage by pandemic response administration staff at the hospital as a stop-gap measure in the event that standard N95 PPE became depleted. The KC Mask was not widely utilized, however, because Truman Medical Center maintained adequate PPE supply chains up to the time of this publication.

## DISCUSSION

The results of Truman Medical Center’s approval of the KC Mask are promising for this N95 stop-gap substitute. Although qualitative fit testing is less rigorous than quantitative fit testing, it has been shown to be highly correlated with proper mask seal and fit.^10^ More extensive testing can and should be done, including quantitative fit testing and design modification.

In light of the current persistent PPE shortages, as well as potential shortages in the case of increasing caseloads, it is important to consider PPE and N95 alternatives, particularly those outside of the main supply chain of conventional PPE. This is critical for a number of reasons, the most obvious being stopgap PPE in health care settings in situations of dire shortages. Additionally, alternative PPE such as that described in this report can create access to PPE for additional groups without compromising current PPE supply channels. This could include providing N95 mask alternatives to individuals living with somebody who tests positive for COVID-19, a situation in which more readily available cloth or surgical masks would not be sufficient. This is especially important when considering that shortages of PPE are particularly common in minority communities and communities of low socio-economic status.^8^ As stated by Brown Emergency Medicine (Rhode Island) physician Dr. Megan Ranney, “Whether it’s the elderly, minority populations, populations affected by structural racism and historical injustice, those are the very groups that have the most difficult time getting adequate PPE for their workers.”^8^ PPE shortages are largely seen as one of the biggest drivers of the racial divide in the COVID-19 pandemic in which infection rates and hospitalization rates disproportionately favor African American and Latino populations.^8^ Accordingly, technology such as that which is presented in this report has the potential to uniquely help our country’s most vulnerable.

Although further analysis and study is needed for this design, persistently increasing caseloads and PPE shortages necessitates an urgent dissemination of these preliminary results. The authors do not advocate for the KC Mask as a replacement of traditional N95 masks or other PPE but do endorse the KC Mask as a stopgap measure, proven to be effective in situations of dire PPE shortage.

## CONCLUSION

The results of Truman Medical Center’s approval of the KC Mask are promising for this N95 alternative. Although further analysis and study is needed for this design, persistently increasing caseloads and PPE shortages necessitates an urgent dissemination of these preliminary results. The authors do not advocate for the KC Mask as a replacement of traditional N95 masks or other PPE but do endorse the KC Mask as a stopgap measure, proven to be effective in situations of dire PPE shortage.

## Data Availability

The sources for all data referenced in this manuscript can be found in the references section of the manuscript.

